# Mitigating Machine Learning Bias Between High Income and Low-Middle Income Countries for Enhanced Model Fairness and Generalizability

**DOI:** 10.1101/2024.02.01.24302010

**Authors:** Jenny Yang, Lei Clifton, Nguyen Thanh Dung, Nguyen Thanh Phong, Lam Minh Yen, Doan Bui Xuan Thy, Andrew A. S. Soltan, Louise Thwaites, David A. Clifton

**Author notes:** These authors jointly supervised this work (Louise Thwaites, David A. Clifton).

## Abstract

Collaborative efforts in artificial intelligence (AI) are increasingly common between high-income countries (HICs) and low-to middle-income countries (LMICs). Given the resource limitations often encountered by LMICs, collaboration becomes crucial for pooling resources, expertise, and knowledge. Despite the apparent advantages, ensuring the fairness and equity of these collaborative models is essential, especially considering the distinct differences between LMIC and HIC hospitals. In this study, we show that collaborative AI approaches can lead to divergent performance outcomes across HIC and LMIC settings, particularly in the presence of data imbalances. Through a real-world COVID-19 screening case study, we demonstrate that implementing algorithmic-level bias mitigation methods significantly improves outcome fairness between HIC and LMIC sites while maintaining high diagnostic sensitivity. We compare our results against previous benchmarks, utilizing datasets from four independent United Kingdom Hospitals and one Vietnamese hospital, representing HIC and LMIC settings, respectively.

## 1 Introduction

Collaborative engagements between high-income countries (HICs) and low-to middleincome countries (LMICs) in the development of artificial intelligence (AI) tools represents concerted efforts to combine resources, expertise, and knowledge. These collaborations involve sharing technological advancements, research findings, and data to jointly create and implement AI tools. The goal is to foster cooperative and inclusive approaches that address shared challenges, promote technological equity, and leverage the strengths of each participant. These collaborative endeavors contribute not only to the advancement of AI technologies but also aim to bridge the digital divide and encourage global inclusivity in the development and utilization of AI tools. While these collaborative efforts offer clear benefits, ensuring the fairness and equity of these data-driven tools is crucial, especially considering the unique contexts and challenges faced by LMIC hospitals in comparison to HIC hospitals.

Hospitals in LMICs often contend with resource constraints, including inadequate funding, outdated infrastructure, a shortage of technical expertise, and a limited availability of comprehensive and digitized healthcare data [1–5]—essential requirements for the development and validation of AI algorithms. This often results in a significant disparity in resource availability between HIC and LMIC settings. Notably, the discrepancy in data availability creates a bias during the development and training of collaborative models, affecting their relative effectiveness when deployed across diverse settings, especially where there are substantial variations in socioeconomic status [2, 4, 6, 7]. Consequently, addressing and mitigating unintentional biases in data-driven algorithms is imperative to prevent the perpetuation or exacerbation of existing disparities in healthcare and society.

The presence of bias in machine learning (ML) models, stemming from the composition of data utilized during training, is well-established [8, 9]. Such biases result in divergent performance across specific subgroups in predictive tasks, and hinder a model’s ability to precisely capture the relationship between features and the target outcome. This causes suboptimal generalization and unfair decision-making [8–12]. Previous studies on training fair ML systems has demonstrated the effectiveness of specific ML training frameworks in mitigating biases, particularly those associated with demographic factors. Moreover, our past research demonstrated the effectiveness of these frameworks in successfully mitigating site-specific biases across four distinct hospital groups in the United Kingdom (UK) [8, 9], applied to a COVID-19 screening task. Thus, building upon previous investigations, our present goal is to evaluate the effectiveness of ML debiasing methods across hospitals situated in diverse socioeconomic strata. Through a partnership between the Oxford University Clinical Research Unit (OUCRU) in Ho Chi Minh City, Vietnam, the University of Oxford Institute of Biomedical Engineering in Oxford, England, and the Hospital for Tropical Diseases (HTD) in Ho Chi Minh, Vietnam, our focus specifically extends to hospitals in the UK and Vietnam, representing HIC and LMIC settings, respectively.

In line with insights from [8, 9], our focus will be on two state-of-the-art bias mitigation techniques at the algorithm-level: adversarial debiasing and reinforcement learning (RL) debiasing. To gauge fairness, we will employ the statistical definition of equalized odds [8–11, 13]. Using the same COVID-19 case study, our aim is to mitigate 2 any site-specific biases and assess the effectiveness—considering both classification performance and fairness—of bias mitigation models trained collaboratively across both HIC and LMIC hospital settings. As such, we will utilize datasets from four independent UK hospitals and one Vietnamese hospital, respectively.

## 2 Methods

### 2.1 Datasets

Clinical data comprising linked and deidentified demographic information was obtained from patients across four hospital centers in the UK and one hospital in Vietnam. From the UK, the datasets included electronic health records (EHRs) from hospital emergency departments (EDs) in Oxford University Hospitals NHS Foundation Trust (OUH), University Hospitals Birmingham NHS Trust (UHB), Bedfordshire Hospitals NHS Foundations Trust (BH), and Portsmouth Hospitals University NHS Trust (PUH). These datasets have received approval from the United Kingdom National Health Service (NHS) through the national oversight/regulatory body, the Health Research Authority (HRA), for the development and validation of artificial intelligence models aimed at detecting COVID-19 (CURIAL; NHS HRA IRAS ID: 281832). The data from Vietnam was sourced from the intensive care units (ICUs) in the Hospital for Tropical Diseases (HTD), and approval for its use was obtained from HTD.

The four UK datasets used in training are identical to those utilized in prior studies [9, 14–17], ensuring a consistent approach across investigations. Specifically, for OUH, we encompassed all patients presenting and admitted to the ED. As for PUH, UHB, and BH, the inclusion criteria comprised all patients admitted to the ED. The full inclusion and exclusion criteria for patient cohorts can be found in Supplementary Section B.

From OUH, we obtained three data extracts corresponding to distinct periods: pre-pandemic presentations (Before December 1, 2019), the first wave of the COVID-19 epidemic in the UK (December 1, 2019, to June 30, 2020), and the second wave (October 1, 2020, to March 6, 2021). The positive COVID-19 presentations from “wave one” and pre-pandemic controls were employed in training and continuous validation, with an 80% to 20% random split, respectively.

Similar to earlier investigations [9, 14–17], our focus is on rapid patient triaging, acting as a preliminary measure during the period when confirmatory laboratory testing is awaiting results or when access to definitive molecular testing for COVID-19 is constrained. Consequently, these datasets encompass a segment of regularly acquired clinical data, comprising initial blood tests, vital signs, and the confirmation of COVID-19 diagnosis through a polymerase chain reaction (PCR) swab test (additional details can be found in the subsequent section).

During the initial wave, challenges such as incomplete testing and the imperfect sensitivity of the PCR swab test led to uncertainties in determining the viral status of patients who were either untested or tested negative [16]. To address this, following the methodology used in [9, 14–17], each positive COVID-19 presentation from “wave one” was matched with a set of pre-pandemic negative controls based on age. Using patient 3 presentations from OUH prior to the global COVID-19 outbreak guarantees that these cases are COVID-free. Thus, this careful selection of data ensures the accuracy of COVID-19 status labels used during the training phase of the model. For our purposes, we employed a ratio of 20 controls to 1 positive presentation for the training set. This matching approach aimed to simulate a disease prevalence of 5%, consistent with the actual COVID-19 prevalences observed at all four UK sites during the data extraction period (ranging from 4.27% to 12.2%). It is important to note that this matching process was exclusively applied to the training set, as it directly influences the model weights and biases. The continuous validation set, used solely to evaluate model training and determine an evaluation threshold (without altering the model itself), retains the full stratification without simulating a 5% prevalence. To account for the uncertainty in negative PCR results, a sensitivity analysis was conducted, yielding improvements in the apparent accuracy of the models, as outlined in [8, 14].

The “wave two” dataset, comprising both negative and positive COVID-19 cases confirmed through PCR testing, was designated as the held-out test set.

Since UHB, PUH, BH, and HTD each provided a single extract, we divided each into training, continuous validation, and test sets through a random split (allocated at 60%, 20%, and 20%, respectively). This division was stratified based on the COVID-19 status, which was determined through confirmatory PCR testing. A full summary of each respective dataset can be found in Supplementary Table B1.

As outlined in [3], it’s important to highlight that HTD functions as a specialized hospital primarily dedicated to infectious diseases. Thus, throughout the pandemic, patients experiencing severe COVID-19 or other critical infections were frequently admitted to either the ICU or high dependency units. Noteworthy is the considerable variability in the terminology used for recording COVID-19 cases, encompassing terms such as “COVID-19 lower respiratory infection,” “COVID-19 pneumonia,” “SARS-COV-2 Infection,” “COVID-19 acute respiratory distress syndrome,” “Acute COVID-19,” and various others. Consistent with the approach detailed in [3], any label indicating the presence and severity of COVID-19 was categorized as “COVID-19 positive.”

Finally, we merged data from all sites, yielding final training, continuous validation, and held-out test sets comprising 42,385, 33,371, and 33,095 presentations, respectively (including 2,912, 944, and 2,805 COVID-19 positive cases, respectively). A summary of each dataset is provided in Table 1.

**Table 1.** Aggregate of patients, positive COVID-19 instances, and distribution across hospitals in the training, continuous validation, and test sets.

|  | Training | Continuous Validation | Test |
| --- | --- | --- | --- |
| n, Patients | 42,385 | 33,371 | 33,095 |
| COVID-19 positive (%) | 2,912 (6.9%) | 944 (2.8%) | 2,805 (8.5%) |
| OUH (%) | 11,676 (27.5%) | 23,132 (69.3%) | 22,857 (69.1%) |
| UHB (%) | 6,175 (14.6%) | 2,059 (6.2%) | 2,059 (6.2%) |
| PUH (%) | 22,737 (53.6%) | 7,580 (22.7%) | 7,579 (22.9%) |
| BH (%) | 705 (1.7%) | 236 (0.7%) | 236 (0.7%) |
| HTD (%) | 1,092 (2.6%) | 364 (1.1%) | 364 (1.1%) |

### 2.2 Clinical Features

To ensure a meaningful comparison with prior studies [3, 8, 9, 14–17], we opted for a similar feature set. Prioritizing scalability for rapid triaging, our models were trained on a focused subset of routinely collected clinical data, including initial laboratory blood tests (encompassing full blood counts, liver function tests, and electrolytes), along with vital signs.

It’s important to note that certain features, such as C-reactive protein, bilirubin, albumin, alkaline phosphatase, urea, and estimated glomerular filtration rate (found in the UK datasets), are not part of the standard admission protocol at HTD. Therefore, in order to integrate data from both the UK and HTD, we had to match the features available in the UK hospitals with those present in the HTD database. Additionally, any features with missing values exceeding 30% were excluded. Table 2 summarizes the final features included.

**Table 2.** Clinical predictors considered for COVID-19 diagnosis.

| Category | Matched UK and Vietnam |
| --- | --- |
| Vital Signs | Heart rate, respiratory rate, systolic blood pressure, diastolic blood pressure, temperature |
| Blood Test | Haemoglobin, haematocrit, white cell count, platelets, mean cell volume, neutrophil count, lymphocyte count, monocyte count, eosinophil count, basophil count |
| Liver Function Tests | Alanine aminotransferase |
| Electrolytes | Sodium, potassium, creatinine |

### 2.3 Pre-processing

We first verified uniformity in the units used for identical features. Following this, all features were standardized to have a mean of 0 and a standard deviation of 1. This standardization process aids in achieving convergence in neural network models [18–20]. For handling missing values within the dataset, we employed population median imputation. These pre-processing steps are consistent with established practices in prior studies [3, 8, 9, 14, 15, 17].

### 2.4 Model Architectures

#### XGBoost

In prior research on COVID-19 detection [8, 14, 15], XGBoost demonstrated robust classification performance, establishing itself as a dependable benchmark for this task. Consequently, we initiate the training process with an XGBoost model, employing it as a reference point for evaluating subsequent neural network models. Additionally, we assess feature importance, inherently determined by XGBoost through score assignments during training.

#### Neural Network

Past studies have demonstrated that a conventional fully-connected neural network attains excellent performance for COVID-19 classification [8, 9, 17]. Moreover, a neural network architecture forms the basis for the advanced debiasing methods assessed in this study (namely, an RL-based framework and an adversarial debiasing framework), establishing it as a robust baseline for comparison. In the context of the binary task of COVID-19 classification, the rectified linear unit (ReLU) activation function was used in the hidden layers, and the Sigmoid activation function was used in the output layer of the network.

#### Adversarial Debiasing Model

Adversarial debiasing stands out as a state-of-the-art technique for fairness-aware ML, serving to alleviate and diminish undesired biases in models, particularly those linked to sensitive attributes like gender, race, or other protected characteristics [9, 13, 25, 26]. The primary objective of adversarial debiasing is to improve the fairness of a model’s predictions by minimizing the influence of these sensitive attributes in the decision-making process. This methodology entails training a model with a dual-component structure: a primary task network and a debiasing network. The primary task network focuses on generating desired predictions (in our case, classifying COVID-19 status), while the debiasing network, also referred to as the adversary, seeks to identify and counteract the impact of a sensitive attribute on the model’s predictions (in our case, the hospital where a patient received care). Notably, this technique has proven effective in mitigating bias associated with hospital location across the UK sites examined in this study [9]. We extend this prior work by incorporating data from HTD, a hospital in Vietnam, enabling us to assess bias across diverse socioeconomic strata. Accordingly, we implement the same adversarial debiasing framework as detailed in [9].

#### Reinforcement Learning Debiasing Model

Recently, a novel approach to debiasing, based on reinforcement learning (RL), has emerged as a paradigm shift to standard supervised learning algorithms (the conventional method of tackling classification tasks)[8]. In the RL-based method, an “agent” engages with the input, determining its class and receiving immediate rewards based on that prediction. Positive rewards are granted for correct predictions, while negative rewards are assigned otherwise. This iterative feedback guides the agent in learning the optimal “behavior” to correctly classify samples, maximizing cumulative rewards. The methodology introduced in [8] demonstrates that employing a specialized reward function designed to additionally mitigate unwanted biases can effectively enhance classification fairness with respect to a sensitive attribute. This technique has proven effective in addressing bias related to hospital location across the UK sites examined in this study [8]. Once again, we expand upon this prior work by including data from HTD, enabling the assessment of bias across diverse socioeconomic strata. Consequently, we implement the same RL debiasing framework as outlined in [8].

### 2.5 Experimental Outline

For each task, we employ a training set to select hyperparameters and train the models. Details on the hyperparameter values used in the final models can be found in Supplementary Section C.

A continuous validation set is utilized for ongoing validation and threshold adjustment. It should be noted that for classification tasks, an ML algorithm’s raw output is the probability of class membership, later mapped to a specific class. Therefore, using the continuous validation set, we conduct a grid search to fine-tune the sensitivity/specificity for identifying COVID-19 positive or negative cases. Following the approach of previous studies [8, 9], we opt to optimize the threshold to achieve sensitivities of 0.9 (*±* 0.05). This selected sensitivity exceeds the sensitivity of lateral flow device (LFD) tests, which achieved 56.9% sensitivity for OUH admissions between December 23, 2021, and March 6, 2021 [14]. Additionally, real-time PCR was found to have estimated sensitivities between 80% and 90% [23, 24]. Thus, by optimizing for a sensitivity of 0.9, we ensure clinically acceptable performance in detecting positive COVID-19 cases, comparable to the sensitivities of the current diagnostic gold standard.

After successful development and training, the held-out test set is used to assess the performance of the final models.

To establish a benchmark for comparing our neural network-based models, we start by training an XGBoost model utilizing the complete set of features outlined in Table 2. This model will additionally be used to assess the significance of variables for the classification task.

Following this, we proceed to train a neural network baseline using the entire feature set, facilitating comparison to various bias mitigation methods.

Regarding bias, our initial step involves exploring potential sources of bias across hospital sites. To achieve this, we utilize the Kolmogorov-Smirnov test for assessing covariate shift. Identifying features displaying the most significant distribution shift between the UK sites and HTD, we subsequently exclude these features and train models using a reduced feature set. This approach effectively minimizes the impact of covariate shift in the training data.

For comparison, we then proceed to train new models using the complete feature set, incorporating bias mitigation techniques, namely adversarial debiasing [9, 13, 25, 26] and RL-based debiasing [8].

This thorough comparison enables us to assess the models and techniques that contribute to enhancing classification fairness, thereby reducing undesired bias, in collaborative training across various hospital sites.

### 2.6 Metrics

To assess the performance of the trained models, we present the following metrics: area under the receiver operating characteristic curve (AUROC), area under the precision-recall curve (AUPRC), sensitivity, specificity, positive predictive value (PPV), and negative predictive value (NPV). It should be noted that PPV and NPV are prevalence-dependent, and thus, they are calculated based on the actual prevalence within each test set. These metrics are accompanied by 95% confidence intervals (CIs), computed using 1000 bootstrapped samples drawn from the test set. Tests of significance, indicated by *p*-values, involve evaluating how often one model outperforms another across 1000 pairs of bootstrapped iterations. We use 0.05 as the threshold value for determining statistical significance, unless otherwise stated.

Regarding fairness, our objective is to optimize for equalized odds. In our study, we aim to predict labels *y*_*i*_*ϵ*{0, 1} for samples *i* with features *x*_*i*_. A subgroup of samples *Z* is designated as sensitive (*Z′* is the non-sensitive complement) based on a real-world attribute (in our case, hospital location). Equalized odds, as defined, deems a classifier *Ŷ* fair if *Ŷ* and *Z* are conditionally independent given *Y* [8–11]. For binary classification, this translates to *P*(*Ŷ* = 1|*Y* = *y, Z* = 0) = *P*(*Ŷ* = 1|*Y* = *y, Z* = 1), *yϵ* {0, 1}. In simpler terms, a classifier achieves fairness if true positive rates and false positive rates are equal across all potential classes of the sensitive attribute [8, 9, 27]. When evaluating multiple labels (i.e., when *N*_*Z*_ > 2), we employ the standard deviation (SD) of true positive and false positive scores [8, 9]. SD scores approaching zero indicate greater outcome fairness. As used in [8, 9], the formulas utilized for calculating true positive and false positive SD scores are as follows:

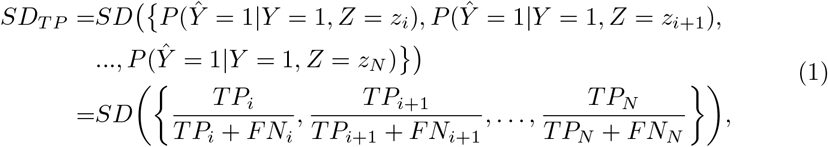

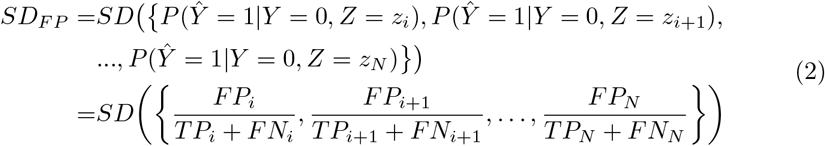

## 3 Results

During the data extraction period, COVID-19 prevalences at all four UK sites varied from 4.27% to 12.2%. The BH cohort exhibited the highest COVID-19 prevalence, attributed to the assessment timeframe covering the second wave of the UK pandemic from January 1, 2021, to March 31, 2021 (12.2% compared to 5.29% in PUH and 4.27% in UHB). As anticipated, the prevalence at HTD was notably higher (74.7%), given its exclusive focus as an infectious disease hospital, managing the most severe cases of COVID-19.

Among all cohorts from the UK and Vietnam, every matched feature exhibited a statistically significant difference in population median (Kruskal-Wallis, *p* < 0.0001). Comprehensive summary statistics, including medians and interquartile ranges, for vital signs and blood tests across all datasets are provided in Supplementary Tables D3 and D4, respectively.

Previously, in studies focused on COVID-19 detection [8, 14, 15], XGBoost exhibited strong classification performance, serving as a reliable benchmark for assessing neural network-based models. When utilizing all features during training, the XGBoost model attained an area under the receiver operating characteristic curve (AUROC) of 0.876 (0.870-0.882) and a sensitivity of 0.801 (0.789-0.813). These results align closely with findings from comparable studies that utilized similar features and patient cohorts, reporting AUROC performances ranging from 0.836 to 0.900 [8, 14, 15].

Regarding fairness, when assessed on the test set, the XGBoost model attained equalized odds (EO) values (represented as standard deviation) of 0.086 and 0.264 for true positive (TP) and false positive (FP) rates, respectively (Table 4).

To evaluate the significance of variables in the classification process, we conducted feature ranking for the XGBoost model by examining the importance scores assigned to each feature during the training phase. Figure 1 depicts the relative importance of the features utilized in the training. Notably, respiratory rate emerged as the most crucial variable, followed by granulocyte counts (specifically basophils and eosinophils) and temperature. This observation aligns with the feature rankings identified in [14, 16] using Shapley Additive Explanations, where both granulocyte counts and respiratory rate were found to be influential features in the classification of COVID-19.

**Figure 1.**
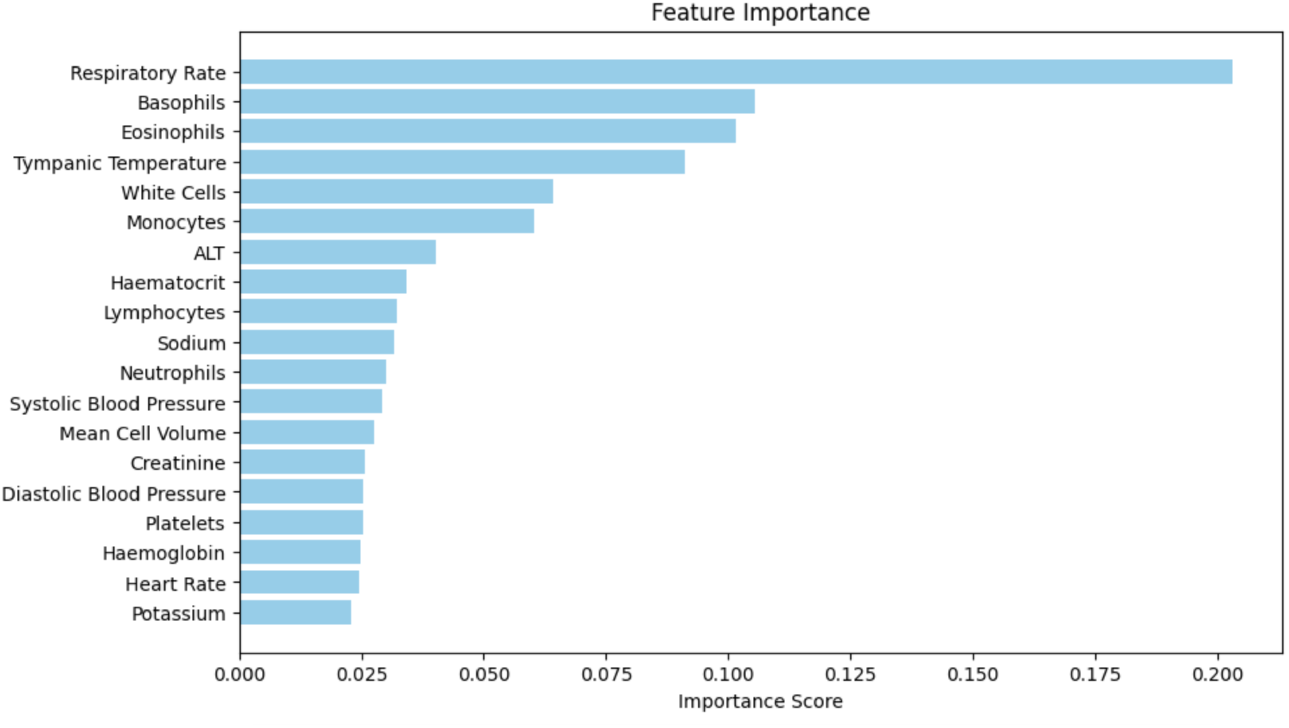
Feature Ranking from XGBoost Model. The bar chart illustrates the feature importance scores obtained from the trained XGBoost model. Each bar represents the relative importance of a specific feature in predicting the target variable. Features with higher importance scores contribute more to the model’s predictive performance.

As a reference point, we then trained a conventional fully-connected neural network (NN) utilizing the complete feature set. This serves as a baseline for assessing the comparative impacts of different bias mitigation techniques. The NN model demonstrated an AUROC of 0.866 (0.86-0.873) and a sensitivity of 0.811 (0.799-0.823) on the test set. This performance is within 1% to that of the XGBoost model, indicating the effectiveness of our initial training in establishing a robust NN model. When comparing the NN model to XGBoost, the difference in performance was found to be statisically significant (*p* < 0.0001 based on 1,000 bootstrapped iterations).

Regarding fairness, the baseline NN model achieved equalized odds values of 0.075 and 0.246 for the TP and FP rates, respectively. These values, slightly lower than those obtained with XGBoost, suggest an enhancement in fairness compared to the XGBoost model.

To investigate potential sources of bias among hospital sites, we analyzed covariate shift using the Kolmogorov-Smirnov (KS) test for two-sample hypothesis testing. Using each of the UK datasets as the reference, we compared the input features between each UK site and the HTD dataset. Our examination revealed respiratory rate and temperature as features displaying the most notable distribution shifts. Here, the KS statistics ranged from 0.636 to 0.804, with *p* < 0.0001 across all UK and HTD pairs (these p-values remained significant after Bonferroni Correction). In contrast, all other features showed KS statistics below 0.5 across all UK and HTD pairs. Notably, when scrutinizing feature distribution differences among solely the UK sites, the KS statistic remained below 0.5 (0 < *p* < 0.677) across all features for all UK hospital pairs. This suggests a more pronounced bias between the UK sites and HTD, compared to biases exclusively between the UK sites.

Consequently, in addition to a model that encompasses all features, we proceeded to train distinct models (without any inherent bias mitigation technique) on feature subsets excluding respiratory rate, excluding temperature, and excluding both features. This approach enables us to compare the performance of models trained on feature sets with reduced covariate shift, to models that include these features, but instead, implement a bias mitigation technique (aimed at addressing bias stemming from different centers).

Training the same neural network (NN) with all features except respiratory rate resulted in a decreased AUROC of 0.839 (95% CI: 0.832-0.847) when compared to the baseline). While sensitivity remained comparable at 0.819 (0.808-0.831), there was a reduction in specificity from 0.761 (0.757-0.765) to 0.674 (0.669-0.678) compared to the baseline NN. In contrast, excluding temperature during training led to a much smaller decline in performance, with the model achieving an AUROC of 0.863 (0.856-0.870) (p = 0.053 when compared to the baseline using 1,000 bootstrapped iterations). In this scenario, there was a slight increase in sensitivity to 0.827 (0.815-0.838), but at the cost of a decrease in specificity to 0.732 (0.728-0.737).

Omitting both respiratory rate and temperature from training resulted in the model achieving an AUROC of 0.837 (0.830-0.845), comparable to the situation where only respiratory rate was excluded. In this instance, sensitivity improved to 0.835 (0.824-0.847). Once again, this heightened sensitivity came at the cost of a reduction in specificity to 0.634 (0.629-0.638). Although the deviation in AUROC from the standard model was statistically significant (*p* < 0.0001), it was found to be statistically similar to the model trained on all features except respiratory rate (*p* = 0.168).

Regarding fairness, models trained with the exclusion of respiratory rate and without both respiratory rate and temperature demonstrated a distinct enhancement, as evidenced by decreased equalized odds for TP and FP rates, ranging between 0.052-0.053 and 0.186-0.196, respectively. When only temperature was excluded from the training set, there was still an improvement in TP and FP equalized odds, albeit to a lesser extent than when respiratory rate was excluded, achieving values of 0.070 and 0.226, respectively.

Despite the apparent improvement in fairness achieved by removing features exhibiting the most bias (in terms of data drift), there was a notable decline in classification performance. As both respiratory rate and temperature were identified as important features during training (according to XGBoost rankings), we proceeded to train two state-of-the-art bias mitigation models - a reinforcement learning (RL) debiasing model and an adversarial debiasing model. These models enable us to leverage all features while mitigating site-specific biases during the training process.

Both RL and adversarial debiasing techniques achieved AUROCs similar to the baseline neural network, with values of 0.858 (0.851-0.864) and 0.852 (0.845-0.859), respectively. Although slightly lower than the baseline NN’s performance (*p* = 0.001 for RL and *p* < 0.0001 for adversarial models), it outperformed all NNs trained on reduced feature sets, except for the NN trained without temperature. When comparing RL and adversarial debiasing models to those trained on reduced feature sets, the differences in performance were found to be statistically significant (0 < *p* < 0.021).

The RL debiasing model exhibited a notable improvement in fairness, with TP and FP equalized odds decreasing to 0.056 and 0.204. These values are akin to the equalized odds observed in models trained with the exclusion of respiratory rate and without both respiratory rate and temperature, although slightly higher. Importantly, this represents a significant enhancement in fairness compared to the baseline NN. Conversely, the adversarial debiasing model only marginally improved upon the baseline, achieving equalized odds of 0.074 and 0.227 for TP and FP standard deviations, respectively.

Full numerical results including AUROC, area under the precision-recall curve (AUPRC), sensitivity, specificity, positive predictive value (PPV), and negative predictive value (NPV) can be found in Table 3.

**Table 3.** COVID-19 status prediction test results across different models. Model performance is optimized to sensitivities of 0.9, and PPV and NPV are reported using true prevalences. Metrics are reported alongside 95% confidence intervals based on 1,000 bootstrapped samples. Bolded values represent the best (†) and second best scores. *RR: Respiratory Rate, T: Temperature.

| Model | AUROC | AUPRC | Sensitivity | Specificity | PPV | NPV |
| --- | --- | --- | --- | --- | --- | --- |
| XGBoost (All features) | **0.876(0.870-0.882)†** | **0.609(0.593-0.623)†** | 0.801(0.789-0.813) | **0.788(0.784-0.792)†** | **0.259(0.254-0.264)†** | 0.977(0.976-0.979) |
| NN (All features) | **0.866(0.860-0.873)** | **0.551(0.534-0.568)** | 0.811(0.799-0.823) | 0.761(0.757-0.765) | 0.239(0.235-0.244) | **0.978(0.976-0.979)** |
| NN (Remove RR) | 0.839(0.832-0.847) | 0.500(0.484-0.517) | 0.819(0.808-0.831) | 0.674(0.669-0.678) | 0.189(0.186-0.192) | 0.976(0.974-0.977) |
| NN (Remove T) | 0.863(0.856-0.870) | 0.538(0.520-0.555) | 0.827(0.815-0.838) | 0.732(0.728-0.737) | 0.222(0.219-0.226) | **0.979(0.977-0.980)†** |
| NN (Remove RR & T) | 0.837(0.830-0.845) | 0.467(0.451-0.485) | **0.835(0.824-0.847)†** | 0.634(0.629-0.638) | 0.174(0.172-0.177) | 0.977(0.975-0.978) |
| RL (All features) | 0.858(0.851-0.864) | 0.517(0.500-0.535) | **0.829(0.817-0.840)** | 0.716(0.712-0.720) | 0.213(0.210-0.216) | **0.978(0.977-0.98)** |
| Adversarial (All features) | 0.852(0.845-0.859) | 0.535(0.518-0.552) | 0.784(0.773-0.796) | **0.773(0.769-0.777)** | **0.242(0.238-0.247)** | 0.975(0.973-0.976) |

When analyzing the subset of test data from HTD, the XGBoost model demonstrated the highest AUROC among all models, achieving a score of 0.836 (0.796-0.873). This figure is slightly lower when compared to the AUROCs achieved across each UK site, ranging from 0.859 to 0.909. In contrast, the standard NN (trained using all features) attained a lower AUROC at HTD than XGBoost, with a score of 0.723 (0.672-0.770) (*p* < 0.0001 when comparing the difference in performance between XG-Boost and the baseline NN). This is notably lower than the AUROCs attained on the UK datasets, which ranged from 0.853 to 0.901. Despite XGBoost’s robust performance at HTD compared to the baseline NN, performance across the UK sites was similar for both models.

Models trained with the exclusion of respiratory rate and without both respiratory rate and temperature exhibited lower AUROC performance at HTD, suggesting a potential decrease in model generalizability and, consequently, increased algorithmic bias between different hospital sites. In this context, the AUROC at HTD was 0.677 (0.622-0.727) and 0.694 (0.640-0.741) for each model, respectively (*p* < 0.0001 and *p* = 0.032 when comparing the difference in performance to the baseline NN, respectively). This contrasts with significantly higher AUROCs ranging from 0.826-0.885 and 0.812-0.881 on the UK datasets, respectively. Models trained without temperature improved in terms of performance on the HTD dataset, relative to the baseline, improving to 0.723 (0.672-0.770) (*p* = 0.008). And, similar to the baseline, AUROCs on the UK datasets ranged from 0.845 to 0.911. Again, despite varying scores at HTD compared to the XGBoost and baseline NN models, performances across the UK sites remained similar across all models.

The RL debiasing model attained the second-highest AUROC on the HTD dataset, achieving a score of 0.781 (0.734-0.824) (*p* < 0.0001 when comparing the difference in performance to the baseline NN and the models trained on reduced feature sets). Once again, this figure is lower than the AUROCs achieved at the UK sites, which ranged from 0.842 to 0.888. Adversarial debiasing similarly reached a high AUROC score of 0.757 (0.711-0.802) on the HTD dataset (*p* = 0.072 when comparing the difference in performance to the baseline NN, and *p* = 0.002 and 0.004 when compared to the models trained on reduced feature sets), in contrast to AUROCs ranging from 0.830 to 0.849 on the UK datasets. When using algorithm-level bias mitigation methods, generalizability across the UK sites and HTD seems to improve compared to models trained using reduced feature sets. As observed previously, while there were varying performances at HTD, performances achieved across the UK sites remained similar across all models.

Full numerical results for HTD including AUROC, AUPRC, sensitivity, specificity, PPV, and NPV can be found in Table 5.

**Table 4.** Equalized odds evaluation for hospital bias across different models. Model performance is optimized to sensitivities of 0.9. Bolded values represent the best (†) and second best scores. *RR: Respiratory Rate, T: Temperature.

| Model | EO (TP) | EO (FP) |
| --- | --- | --- |
| XGBoost (All features) | 0.086 | 0.264 |
| NN (All features) | 0.075 | 0.246 |
| NN (Remove RR) | **0.052** | **0.196†** |
| NN (Remove T) | 0.070 | 0.226 |
| NN (Remove RR and T) | **0.053†** | **0.186** |
| RL (All features) | 0.056 | 0.204 |
| Adversarial (All features) | 0.074 | 0.227 |

**Table 5.** COVID-19 status prediction test results on the HTD subset, across different models. Model performance is optimized to sensitivities of 0.9, and PPV and NPV are reported using true prevalences. Metrics are reported alongside 95% confidence intervals based on 1,000 bootstrapped samples. Bolded values represent the best (†) and second best scores. *RR: Respiratory Rate, T: Temperature.

| Model | AUROC | AUPRC | Sensitivity | Specificity | PPV | NPV |
| --- | --- | --- | --- | --- | --- | --- |
| XGBoost (All features) | **0.836(0.796-0.873)†** | **0.944(0.926-0.960)†** | **0.989(0.978-0.997)†** | 0.159(0.099-0.226) | 0.787(0.775-0.800) | **0.824(0.660-0.956)†** |
| NN (All features) | 0.723(0.672-0.770) | 0.887(0.855-0.917) | **0.971(0.953-0.986)** | 0.170(0.105-0.240) | 0.786(0.773-0.801) | **0.652(0.481-0.810)** |
| NN (Remove RR) | 0.694(0.640-0.741) | 0.880(0.848-0.910) | 0.931(0.904-0.956) | 0.205(0.141-0.280) | 0.786(0.771-0.803) | 0.486(0.360-0.617) |
| NN (Remove T) | 0.768(0.721-0.809) | **0.913(0.889-0.937)** | 0.957(0.936-0.975) | 0.216(0.147-0.288) | **0.793(0.778-0.809)** | 0.613(0.468-0.742) |
| NN (Remove RR & T) | 0.677(0.622-0.727) | 0.867(0.831-0.900) | 0.917(0.888-0.940) | 0.216(0.146-0.290) | 0.786(0.770-0.802) | 0.452(0.333-0.569) |
| RL (All features) | **0.781(0.734-0.824)** | **0.913(0.885-0.940)** | 0.942(0.917-0.963) | **0.239(0.170-0.313)†** | **0.795(0.780-0.812)†** | 0.568(0.439-0.685) |
| Adversarial (All features) | 0.757(0.711-0.802) | 0.904(0.872-0.932) | 0.938(0.913-0.960) | **0.227(0.155-0.307)** | 0.792(0.777-0.810) | 0.541(0.412-0.665) |

## 4 Discussion

Ensuring impactful collaborative AI development that benefits both HIC and LMIC hospitals requires strategies aimed at mitigating inadvertent site-specific biases. With 12 a specific focus on biomedical engineering and AI, our goal was to evaluate the fairness and generalizability of algorithms, especially when deploying a collaboratively-trained model across HIC and LMIC hospitals. We aimed to demonstrate that implementing bias mitigation techniques enhances both algorithmic fairness and generalizability while maintaining effective classification performance. This is particularly crucial in healthcare settings where algorithmic findings directly impact clinical decision-making and patient care. Hence, the objective of this study was to introduce methods aimed at cultivating increased trust among clinicians and patients in the effectiveness and reliability of ML-based technologies. This, in turn, serves to encourage and enhance international collaboration and development initiatives in AI.

In general, we observed that models incorporating some form of bias mitigation, whether through the removal of biased features or through the inclusion of bias mitigation training methods, exhibited greater fairness (with respect to equalized odds) compared to those without such considerations. It’s important to note that this reduction in bias came at a modest cost to performance, as both the removal of features from the training set and the implementation of bias mitigation at the training-level resulted in a decline in performance. The act of excluding features from training removes potentially valuable information that the model could learn from, emphasizing the need to strike a balance when constructing models with the dual objectives of mitigating undesirable biases and training a robust classifier.

We observed that excluding respiratory rate from model training resulted in a more pronounced decrease in AUROC (compared to the baseline model trained on all features) than a model trained without temperature. This aligns with respiratory rate being identified as the most influential feature, relative to all other features, in determining the presence of COVID-19 (as illustrated in Figure 1). Therefore, omitting this feature from model development would have the greatest impact on test performance. This is reinforced by the similarity in performance between the model trained without respiratory rate and the model trained without both respiratory rate and temperature, with further removal of temperature showing no significant impact.

Similarly, as respiratory rate emerged as one of the most biased features (exhibiting greatest data drift) between the UK hospital sites and HTD in Vietnam, excluding this feature from training resulted in improved fairness, as evidenced by noticeable enhancements in both TP and FP equalized odds. In contrast, removing temperature from training led to only a slight improvement in fairness. Despite temperature being identified as highly biased across different sites, its influence on classification performance was comparatively lower than that of respiratory rate. Consequently, its presence or removal had a lesser impact on equalized odds.

Generally, discovering respiratory rate and temperature as the most biased features between the UK hospitals and HTD is not unexpected. Given that HTD is a specialized hospital receiving referrals from other medical facilities, its high workload is an important factor to consider. Consequently, the meticulous counting of respiratory rate may not have been consistently performed unless a patient appeared unwell. In busy LMIC wards, reduced staffing time may also contribute to less precise counting. Similarly, the disparity in temperature readings may be attributed to monitoring differences, as HTD utilizes auxillary non-digital thermometers, resulting in less detailed 13 output. Nonetheless, temperature measurements can exhibit significant variability in any case, thus using a single temperature measurement may be an unreliable indicator [28]. Subsequent experiments may explore the option of excluding temperature as a variable if it proves to be inconsistent, exhibits substantial data shift across different sites, or is determined to have limited impact on the classification.

Regarding algorithmic-level bias mitigation methods, the decrease in performance, as measured by AUROC, was notably less compared to the direct removal of biased features; however, there was still a slight performance decrease. Similar findings have been reported in prior studies exploring bias mitigation methods, where improvements in fairness were achieved at the expense of a marginal reduction in performance [8, 21, 22].

The RL debiasing method significantly enhanced equalized odds, achieving a level comparable to models trained on reduced feature sets, while maintaining robust classification performance (similar to standard models trained on all features). In contrast, although adversarial debiasing also upheld strong classification performance, its impact on equalized odds improvement was comparatively modest. This difference may be attributed to the standard supervised learning setting employed in adversarial debiasing, where cross-entropy loss provides a learning signal irrespective of the presented data. Consequently, a model can become skewed or biased based on the majority class within the batch, due to the aggregation of errors. By opting for an RL setup (instead of a supervised learning framework dependent on gradient descent), one gains control over when and how a learning signal is backpropagated, thereby reducing the risk of skewing a model towards the majority class [8]. In our case, this approach mitigated the risk of biasing the model towards the distribution from the location with the most data points in training, i.e., the UK. Nevertheless, since there is no universal solution applicable to all datasets and machine learning scenarios, both options (among others) can be evaluated for different tasks.

We found that generalizability, measured by AUROC, was optimal when using the XGBoost and baseline NN models. However, corresponding fairness metrics were the poorest among all models. It is crucial to highlight that the equalized odds metrics used in the evaluation are based on TP and FP rates, determined after thresholding. Consequently, despite a high AUROC, the model exhibited bias toward the distributions in the UK datasets, resulting in the classification threshold performing suboptimally on the HTD dataset.

Moreover, the removal of features with the most distribution drift also led to a significant decrease in generalizability. This could be attributed to the fact that these features were identified as highly influential in accurately classifying COVID-19. Therefore, their removal made it more challenging to correctly classify patients, especially those from an external hospital site or distribution.

On the other hand, with the application of bias mitigation techniques, generalizability increased, as evidenced by the improved AUROC on the HTD dataset compared to both the baseline NN and NN models trained on a reduced number of features. However, the NN trained on all features except temperature slightly outperformed the adversarial debiasing model, which again, may relate to the trade-off between fairness 14 and accuracy [8, 21, 22]. Although these performances did not match those of XG-Boost, fairness, as measured by equalized odds, significantly improved. The focus of bias mitigation methods on reducing bias between different hospital sites likely contributed to increased classification accuracy at HTD, making the algorithm less biased toward the UK sites, despite the majority of the data originating from there.

Although achieving widespread generalizability is desirable for scalability, costeffectiveness, and relevance to diverse cohorts and environments, it is often unattainable. This limitation became evident when comparing performance on the HTD dataset with that on the UK datasets. When conducting subset analysis for each site independently, we noted that AUROCs across all UK datasets were consistently within a similar range across all tested models. However, there was significant variability in performance on the HTD dataset. Despite improvements in performance at HTD through the application of bias mitigation techniques, the AUROC remained significantly lower than the scores achieved on the UK hospital datasets. This is likely due to the fact that the HTD dataset represents the sole LMIC hospital, whereas the UK datasets are all part of the NHS, sharing more similarities with each other. Factors contributing to this divergence between settings may include concept drift in disease patterns (alterations in presentation, prevalence, and characteristics), population variability (patients at one center may not represent those in another location), evolving medical practices (changes in diagnostic, treatment, and management methods), and data drift (changes in patient behaviors, trends, or data collection methods) [1, 3–6, 8, 9]. Moreover, the majority of the data originates from the UK sites. Consequently, despite efforts to mitigate bias, the model remains more attuned to the characteristics of the UK datasets, resulting in better performance on these specific subsets.

Furthermore, HTD, being a specialized hospital for infectious diseases, typically included COVID-19 negative cases involving other infectious diseases, and critical cases with various comorbidities were treated there. Additionally, HTD was specifically designated as a “COVID-19” hospital during the pandemic, primarily receiving referrals for severe COVID-19 cases [3]. Given that the Vietnamese dataset predominantly consisted of severely ill patients, models may face difficulties in accurately distinguishing

COVID-19 from other diseases based on vital signs and blood test features. This challenge arises because other diseases, including infectious ones, may coexist. And, in the context of UK hospitals, a broader spectrum of COVID-19 case severity is evident, covering all individuals presenting to the hospital, with only a small subset progressing to ICUs. As a result, the AI-based diagnosis of COVID-19 becomes a notably more challenging task at HTD, as the model must discern the specific reason for ICU admission, especially in cases involving other infectious diseases. For instance, as highlighted in [3], distinguishing COVID-19 from pneumonia (which is common at HTD, with much more similar clinical features to COVID-19) is more challenging than distinguishing it from cases such as a fractured leg.

This difficulty might also be a factor in the lower observed specificity in the HTD dataset compared to the UK datasets. Therefore, despite a high AUROC at external sites, it may be crucial to tailor the classification threshold (i.e., the criterion for categorizing COVID-19 status as positive or negative) independently for each site to uphold desired levels of sensitivity and specificity [17]. Hence, future studies should

15 delve into identifying an optimal decision threshold, as it directly impacts performance and fairness metrics by altering true positive/true negative rates [9]. Particularly in clinical contexts, maintaining consistent sensitivity/specificity scores across various hospitals is typically desirable, as fluctuations in these metrics may impede clinicians’ trust in a model’s performance [17].

## Contributions

JY conceived and ran the experiments, wrote and implemented the code, and wrote the initial manuscript draft. LC helped advise statistical tests and performance evaluation. LT and JY applied for ethical approval and use of the Vietnam (HTD) data. AAS applied for the ethical approval for the UK (OUH, PUH, UHB, BH) data. JY preprocessed the Vietnam dataset. JY and AAS preprocessed the UK COVID-19 datasets. All authors revised the manuscript.

## Acknowledgements

We express our sincere thanks to all patients and staff across the four participating NHS trusts (Oxford University Hospitals NHS Foundation Trust, University Hospitals Birmingham NHS Trust, Bedfordshire Hospitals NHS Foundations Trust, and Portsmouth Hospitals University NHS Trust) and the Hospital for Tropical Diseases in Vietnam. We also express our thanks to the Critical Care Asia Africa Registry team.

## Ethics

United Kingdom National Health Service (NHS) approval via the national oversight/regulatory body, the Health Research Authority (HRA), has been granted for use of routinely collected clinical data to develop and validate artificial intelligence 16 models to detect Covid-19 (CURIAL; NHS HRA IRAS ID: 281832). The ethics committee at the Hospital for Tropical Diseases (HTD) approved use of the HTD dataset for COVID-19 diagnosis.

## Data Availability

Data from OUH studied here are available from the Infections in Oxfordshire Research Database (https://oxfordbrc.nihr.ac.uk/research-themes/modernising-medical-microbiology-and-big-infection-diagnostics/infections-in-oxfordshire-research-database-iord/), subject to an application meeting the ethical and governance requirements of the Database. Data from UHB, PUH and BH are available on reasonable request to the respective trusts, subject to HRA requirements.

Data from HTD is available from the CCAA Vietnam Data Access Committee, subject to an application meeting the ethical and governance requirements.

## Code Availability

The code used for the adversarial debiasing model is available at https://github.com/yangjenny/adversarial_learning_bias_mitigation/. The code used for the reinforcement learning debiasing model is available at https://github.com/yangjenny/BiasMitigationRL.

