## Supplementary Material for "Mitigating Machine Learning Bias Between High Income and Low-Middle Income Countries for Enhanced Model Fairness and Generalizability"

### Appendix A Software and Implementation

Experiments were performed using Python (v3.8.3). All models were run using an Intel Xeon E-2146G Processor (CPU: 6 cores, 4.50 GHz max frequency). Statistical tests were performed using the SciPy (Statistical Functions) package (v1.10.1). Scikit Learn (v1.3.2) was used for standardization, median imputation, and calculating performance metrics. Performance metrics were calculated using Scikit Learn and manually programmed.

The XGBoost classifier was implemented using the XGBoost package (v2.0.3). Neural network models were implemented using Pytorch (v2.1.2+cu121). The code used for the adversarial debiasing model is available at https://github.com/yangjenny/adversarial_learning_bias_mitigation. Adversarial debiasing was performed using Pytorch (v2.1.2+cu121). The code used for the reinforcement learning debiasing model is available at https://github.com/yangjenny/BiasMitigationRL. Reinforcement learning was performed using Tensorflow (v2.13.1).

### Appendix B Datasets

The following inclusions and exclusions are reproduced from previous studies (Soltan et al., 2022 and Yang et al., 2022a, 2022b, 2023a, and 2023b).

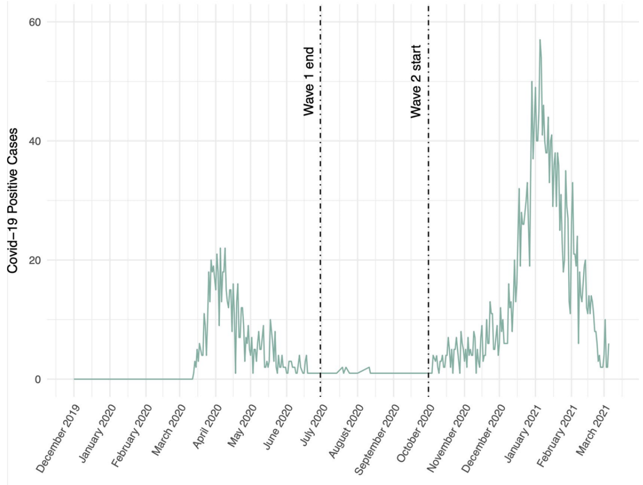


**Supplementary Figure B1** Daily number of patients presenting to Oxford University Hospitals NHS Foundation Trust testing positive for COVID-19, between 1st December 2019 and 6th March 2021.

**Oxford University Hospitals NHS Foundation Trust (OUH):** We included all patients attending acute and emergency care settings at OUH who received routine blood tests on arrival, considering presentations before December 1, 2019, and thus before the pandemic, as the COVID-19-negative (control) cohort. We considered presentations during the 'first wave' of the UK COVID-19 pandemic (December 1,

2019 to June 30, 2020) with PCR confirmed SARS-CoV-2 infection as the COVID-19-positive (cases) cohort. We excluded patients who opted out of electronic health record (EHR) research and those who did not receive laboratory blood tests or were younger than 18 years of age. Due to incomplete penetrance of testing during the first wave of the pandemic, and imperfect sensitivity of the PCR test, there is uncertainty in the viral status of patients presenting during the pandemic who were untested or tested negative. We therefore selected a pre-pandemic control cohort during training to ensure absence of disease in patients labelled as COVID-19-negative. Clinical features extracted for each presentation included first-performed blood tests, blood gases, vital signs measurements and PCR testing for SARS-CoV-2 (Abbott Architect [Abbott, Maidenhead, UK], TaqPath [Thermo Fisher Scientific, Massachusetts, USA] and Public Health England-designed RNA-dependent RNA polymerase assays).

**Portsmouth Hospitals University NHS Foundation Trust (PUH):** PUH considered all patients admitted to the Queen Alexandria Hospital, serving a population of 675,000 and offering tertiary referral services to the surrounding region, between March 1, 2020 and February 28, 2021. Confirmatory COVID-19 testing was by laboratory SARS-CoV2 RT-PCR assay, considering any positive PCR result within 48hrs of admission as a true positive.

**University Hospitals Birmingham NHS Foundation Trust (UHB):** UHB considered all patients admitted to The Queen Elizabeth Hospital, Birmingham, between December 01, 2019 and October 29, 2020. The Queen Elizabeth Hospital is a large tertiary referral unit within the UHB group which provides healthcare services for a population of 2.2 million across the West Midlands. Confirmatory COVID-19 testing was performed by laboratory SARS-CoV-2 RT-PCR assay.

**Bedfordshire NHS Foundation Trust (BH):** BH considered all patients admitted to Bedford Hospital between January 1, 2021 and March 31, 2021. BH provides healthcare services for a population of around 620,000 in Bedfordshire. Confirmatory COVID-19 testing was performed on the day of admission by point-of-care PCR based nucleic acid testing [SAMBA-II & Panther Fusion System, Diagnostics in the Real World, UK, and Hologic, USA].

**Hospital for Tropical Diseases (HTD):** HTD considered all patients admitted between January 1, 2021 and December 31,2022.

**Supplementary Table B1** Total patients and positive COVID-19 cases in the OUH (OUH pre-pandemic, "wave one" and "wave two"), UHB, PUH, BH, and HTD datasets. Note, the number shown for the OUH pre-pandemic dataset is the total before matching controls.

| | Cohort | Total Patients | COVID-19 Positive Cases |
|---|---|---|---|
| OUH pre-pandemic | Before Dec 1/19 | 114,957 | 0 |
| OUH "wave 1" | Dec 1/19-June 30/20 | 701 | 701 |
| OUH "wave 2" | Oct 1/20-Mar 6/21 | 22,857 | 2,012 (8.80%) |
| UHB | Dec 1/19-Oct 29/20 | 10,293 | 439 (4.27%) |
| PUH | Mar 1/20-Feb 28/21 | 37,896 | 2,005 (5.29%) |
| BH | Jan 1/21-Mar 31/21 | 1,177 | 144 (12.2%) |
| HTD | Dec 10/20-Dec 30/22 | 1,820 | 1,360 (74.7%) |

### Appendix C Model Architectures

**Supplementary Table C2** Hyperparameter values used in COVID-19 status prediction task.

| | Hyperparameters: |
|---|---|
| RL | |
| | Gamma = 0.1 |
| | Learning Rate = 0.0001 |
| | Epsilon range = [0.01,1] |
| | Hidden nodes = 400 |
| ADV | |
| | Hidden nodes (predictor) = 100 |
| | Hidden nodes (adversary) = 100 |
| | Dropout = 0.3 |
| | Alpha = 1 |
| | Learning Rate = 0.0001 |
| NN | |
| | Hidden nodes = 10 |
| | Learning rate = 0.05 |
| | Optimizer = Adam |
| XGB | |
| | Learning rate = 0.1 |
| | N estimators = 100 |
| | Depth = 3 |

### Appendix D Additional Results

**Supplementary Table D3** Distribution of vital signs, reported as median and interquartile ranges, for each patient cohort.

| | Oxford University Hospitals (pre-pandemic & wave 1 cases, to June 30/20) | | Oxford University Hospitals | Portsmouth Hospitals University NHS Trust | University Hospitals Birmingham NHS Foundation Trust | Bedfordshire Hospitals NHS Foundation Trust | Hospital for Tropical Diseases | Kruskal-Wallis, p-value |
|---|---|---|---|---|---|---|---|---|
| | Prepandemic cohort | COVID-19-cases cohort | Oct 1/20-Mar 6/21 | Mar 1/20-Feb 28/21 | Dec 1/19 - Oct 29/20 | Jan 1/21-Mar 31/21 | Jan 1/21-Dec 31/22 | |
| Respiratory Rate (breath/min) | 18.0 (16.0-19.0) | 20.0 (18.0-24.0) | 18.0 (16.6-19.0) | 17.0 (16.0-19.0) | 18.0 (17.0-20.0) | 18.0 (16.0-20.0) | 24.0(20.0-26.5) | <0.0001 |
| Heart Rate (beats/min) | 82.0 (71.0-96.0) | 88.0 (75.0-101.0) | 84.0 (72.0-97.0) | 82.0 (71.0-95.0) | 86.0 (73.0-101.0) | 84.0 (73.0-97.0) | 94.0(84.0-108.0) | <0.0001 |
| Systolic Blood Pressure (mmHg) | 132.0 (118.0-150.0) | 131.0 (115.0-146.0) | 134.0 (119.0-152.0) | 128.0 (114.0-146.0) | 136.0 (119.0-155.0) | 131.0 (116.0-149.0) | 130.0(111.5-140.0) | <0.0001 |
| Diastolic Blood Pressure (mmHg) | 74.0 (65.0-84.0) | 74.0 (64.0-84.0) | 75.0 (65.0-85.0) | 76.0 (67.0-84.0) | 77.0 (68.0-87.0) | 78.0 (68.0-88.0) | 80.0(70.0-80.0) | <0.0001 |
| Temperature (C) | 36.5 (36.1-36.9) | 36.9 (36.3-37.6) | 36.3 (36.0-36.7) | 36.3 (36.0-36.8) | 36.7 (36.4-37.2) | 36.5 (36.4-36.9) | 37.0(37.0-37.3) | <0.0001 |

**Supplementary Table D4** Distribution of blood test features, reported as median and interquartile ranges, for each patient cohort.

| | Oxford University Hospitals (pre-pandemic & wave 1 cases, to June 30/20) | | Oxford University Hospitals | Portsmouth Hospitals University NHS Trust | University Hospitals Birmingham NHS Foundation Trust | Bedfordshire Hospitals NHS Foundation Trust | Hospital for Tropical Diseases | Kruskal-Wallis, p-value |
|---|---|---|---|---|---|---|---|---|
| | Prepandemic cohort | COVID-19-cases cohort | Oct 1/20-Mar 6/21 | Mar 1/20-Feb 28/21 | Dec 1/19 - Oct 29/20 | Jan 1/21-Mar 31/21 | Jan 1/21-Dec 31/22 | |
| HAEMOGLOBIN (g/L) | 130.0 (116.0-142.0) | 130.0 (114.0-144.0) | 129.0 (114.0-142.0) | 129.0 (114.0-143.0) | 127.0 (113.0-140.0) | 134.0 (119.0-146.0) | 128.0(113.0-141.0) | <0.0001 |
| WHITE CELLS ($10^9 l^{-1}$) | 8.45 (6.46-11.18) | 6.98 (5.14-9.72) | 8.94 (6.7-12.06) | 8.6 (6.7-11.3) | 9.4 (7.1-12.6) | 9.2 (6.9-12.5) | 9.55(6.69-13.565) | <0.0001 |
| PLATELETS ($10^9 l^{-1}$) | 249.0 (199.0-307.0) | 215.0 (163.0-283.5) | 251.0 (198.0-314.0) | 251.0 (199.0-312.0) | 247.0 (196.0-311.0) | 246.0 (196.0-310.0) | 216.0(152.0-281.0) | <0.0001 |
| HAEMATOCRIT | 0.39 (0.35-0.42) | 0.4 (0.35-0.44) | 0.39 (0.35-0.43) | 0.39 (0.34-0.42) | 0.38 (0.34-0.42) | 0.39 (0.35-0.43) | 0.392(0.35-0.428) | <0.0001 |
| SODIUM (mM) | 138.0 (136.0-140.0) | 136.0 (134.0-139.0) | 138.0 (135.0-140.0) | 138.0 (136.0-140.0) | 137.0 (134.0-139.0) | 138.0 (136.0-140.0) | 134.0(130.0-137.0) | <0.0001 |
| CREATININE (umol/L) | 73.0 (60.0-93.0) | 79.0 (65.0-106.0) | 74.0 (60.0-97.0) | 74.0 (60.0-96.0) | 78.0 (62.0-105.0) | 80.5 (65.75-104.0) | 76.0(61.0-99.0) | <0.0001 |
| POTASSIUM (mM) | 4.0 (3.7-4.3) | 4.0 (3.7-4.3) | 4.0 (3.8-4.4) | 4.2 (3.9-4.4) | 4.1 (3.8-4.4) | 4.3 (4.0-4.6) | 3.72(3.35-4.09) | <0.0001 |
| MEAN CELL VOL (fl) | 89.6 (86.0-93.4) | 90.2 (86.6-94.2) | 90.2 (86.6-94.2) | 89.0 (84.9-93.0) | 89.9 (86.2-93.6) | 88.0 (85.0-92.0) | 89.75(85.675-93.4) | <0.0001 |
| NEUTROPHILS ($10^9 l^{-1}$) | 5.72 (3.99-8.36) | 5.11 (3.48-7.49) | 6.44 (4.4-9.55) | 5.9 (4.2-8.6) | 6.9 (4.7-10.0) | 6.8 (4.7-9.73) | 7.705(4.92-11.3) | <0.0001 |
| LYMPHOCYTES ($10^9 l^{-1}$) | 1.51 (1.0-2.13) | 0.96 (0.65-1.38) | 1.31 (0.85-1.89) | 1.5 (0.97-2.2) | 1.3 (0.9-1.9) | 1.27 (0.86-1.83) | 0.93(0.57-1.54) | <0.0001 |
| MONOCYTES ($10^9 l^{-1}$) | 0.64 (0.48-0.85) | 0.49 (0.35-0.74) | 0.66 (0.48-0.89) | 0.63 (0.48-0.85) | 0.7 (0.5-0.9) | 0.66 (0.48-0.92) | 0.46(0.28-0.69) | <0.0001 |
| EOSINOPHILS ($10^9 l^{-1}$) | 0.1 (0.04-0.2) | 0.01 (0.0-0.06) | 0.07 (0.02-0.16) | 0.1 (0.02-0.2) | 0.1 (0.0-0.2) | 0.06 (0.02-0.16) | 0.02(0.0-0.08) | <0.0001 |
| BASOPHILS ($10^9 l^{-1}$) | 0.04 (0.03-0.06) | 0.02 (0.01-0.03) | 0.04 (0.02-0.06) | 0.04 (0.02-0.06) | 0.1 (0.0-0.1) | 0.05 (0.03-0.07) | 0.01(0.01-0.03) | <0.0001 |
| ALT (IU/L) | 18.0 (13.0-28.0) | 25.0 (17.0-41.0) | 20.0 (13.0-33.0) | 19.0 (13.0-30.0) | 19.0 (13.0-30.0) | 20.0 (13.0-31.0) | 33.0(20.0-60.0) | <0.0001 |
